# Risk of severe COVID-19 in patients with inflammatory rheumatic diseases treated with immunosuppressive therapy in Scotland

**DOI:** 10.1101/2022.02.13.22270898

**Authors:** Paul M McKeigue, Duncan Porter, Rosemary J Hollick, Stuart H Ralston, David A McAllister, Helen M Colhoun

## Abstract

**Objectives:** To investigate the association of severe COVID-19 in those with inflammatory rheumatic diseases (IRD) treated with immunosuppressive drugs.

**Methods:** A list of 4633 patients on biologics and targeted synthetic (ts) DMARDs in March 2020 was linked to a case-control study that includes all cases of COVID-19 in Scotland.

**Results:** By 22 November 2021 433 of the 4633 patients treated with biologics and tsDMARDs had been diagnosed with COVID-19, of whom 58 had been hospitalised. With all those in the population not on DMARDs as reference category, the rate ratio for hospitalised COVID-19 associated with DMARD treatment was 2.14 (95% CI 2.02 to 2.26) in those on conventional synthetic (cs) DMARDs, 2.01 (95% CI 1.38 to 2.91) in those on TNF inhibitors as the only biologic agent, and 3.83 (95% CI 2.65 to 5.56) in those on other biologic agents. Among those on csDMARDs, rate ratios for hospitalised COVID-19 were lowest at 1.66 (95% CI 1.51 to 1.82) in those on methotrexate and highest at 5.4 (95% CI 4.4 to 6.7) in those on glucocorticoids at average dose >10 mg/day prednisolone equivalent.

**Conclusion:** The risk of hospitalised COVID-19 is elevated in IRD patients treated with immunosuppressive drugs. Of these drugs, methotrexate, hydroxychloroquine, and TNF inhibitors carry the lowest risk, JAK inhibitors and B-cell depleting agents a higher risk and prednisolone the highest risk. A larger study is needed to estimate reliably the risks associated with each class of biologic agent.

**Key messages:**

- Risk of hospitalised COVID-19 is about twofold higher in IRD patients treated with immunosuppressive therapies – csDMARDS or biologics – than in the general population.
- Risk is lowest in those treated with methotrexate, hydroxychoroquine and TNF inhibitors. Of the other biologic drugs, treatment with B cell depleters and JAK inhibitors is associated with higher risk but the numbers are too small for risk associated with each drug class to be estimated reliably.
- The risk of severe COVID-19 with glucocorticoids at a dose greater than 10 mg/day prednisolone equivalent is higher than that of any other drug class studied.

## Introduction

Early in the COVID-19 epidemic in the UK, people on “immunosuppression therapies sufficient to increase risk of infection” were designated by public health agencies as clinically extremely vulnerable and thus eligible for shielding [1]. In Scotland letters advising these individuals to shield themselves were issued from April 2020 onwards, modiand in November 2020, a further letter was issued with “extra protection level advice for people at highest risk” based on the current classification of the protection level of the area that they were resident in. The list of those eligible for shielding has been regularly updated, and was used to identify those at highest priority for vaccination. More recently the Joint Committee on Vaccination and Immunisation has recommended a third primary dose of vaccine to achieve maximal protection of those on biologic immunosuppressants, targeted synthetic immunosuppressants, or non-biological oral immune modulating drugs including corticosteroids at dose equivalent to 10mg prednisolone per day [2]. A recent review however concluded that “a diagnosis of inflammatory arthritis, psoriasis, or inflammatory bowel diseases does not increase risk for SARS-CoV-2 infection or severe COVID-19” and that cytokine inhibitors “might even lower the risk of severe COVID-19” [3]. The objective of this study was to investigate the associations of hospitalized or fatal COVID-19 with autoimmune rheumatologic disease and disease-modifying antirheumatic drugs (DMARDs).

## Methods

Because biologic and targeted synthetic immunosuppressants are prescribed only in hospital and not captured by prescribing databases, Public Health Scotland (PHS) requested clinicians in relevant specialties to provide lists of patients on these drugs in March 2020 at the outset of the epidemic [4,5]. This study focuses on lists provided by NHS Lothian, NHS Grampian and part of NHS Greater Glasgow and Clyde covering about one-third of the Scottish population. These lists were not updated. The merged list, hereafter the “biologics list” was linked to the REACT-SCOT case-control study to take advantage of data linkages. This study includes all 700022 diagnosed cases of COVID-19 in Scotland since the start of the epidemic, and 3238432 individuals who have been sampled at least once as controls.

The design of the REACT-SCOT study has been described previously [6]. In brief, for every incident case of COVID-19 in the population ten controls matched for one-year age, sex and primary care practice and alive on the day of presentation of the case that they were matched to were selected using the Community Health Index database. With this incidence density sampling design it is possible and correct for an individual to appear more than once as a control and subsequently as a case. COVID-19 cases are those with a positive nucleic acid test, a hospital discharge diagnosis or death with COVID-19 mentioned on death certificate. The REACT-SCOT case-control dataset is linked to national data on vaccinations, hospital discharges and outpatient consultations in the last 5 years before presentation date, dispensed prescriptions written in primary care in the last 240 days, and the list of individuals eligible for shielding. The dataset used for this study was based on cases presenting up to 22 November 2021 and their matched controls. The main outcome measure was hospitalisation within 14 days of a positive test for COVID-19, or fatal outcome defined as death within 28 days of a positive test or any death certified with COVID-19 as underlying cause. For brevity we refer to this outcome as “hospitalised COVID-19”. Ascertainment of this outcome is complete, as PCR tests are done on all patients admitted to hospital. As only a fraction of SARS-CoV-2 infections in the population are detected by unscheduled testing, and testing rates vary widely, this design cannot be used to study effect on infectionr rates; this would require follow-up of a cohort tested at least ever two weeks.

Patients attending rheumatology clinics were identified from the outpatient specialty codes in the SMR00 (Scottish Morbidity Record) dataset. Outpatient diagnoses are not recorded in SMR. ICD-10 diagnostic codes (as main condition or other condition) for autoimmune rheumatic diseases in hospital discharge records were grouped into three broad categories: rheumatoid arthritis (M05 to M09, M12.3, M13); psoriatic or other seronegative arthritis (M07, M45, M46); and connective tissue disorders (M30 to M35). From drug prescribing records, patients who had received conventional synthetic DMARDs – methotrexate, hydroxychloroquine, sulfasalazine, leflunomide or prednisolone – in the last 240 days were identified. To exclude those who had been prescribed these drugs for non-rheumatic conditions, patients were classified as on csDMARDs only if they had a rheumatology outpatient consultation (specialty code AR) in the last 5 years.

Average weekly dose of prednisolone equivalent was calculated from the total quantity dispensed during the last 120 days before date of presentation as the sum of equivalent doses of prednisolone, cortisone acetate, deflazacort, dexamethasone, methylprednisolone and prednisone [7].

Rate ratios for hospitalised / fatal COVID-19 were estimated by conditional logistic regression, with vaccination status (0, 1 or 2 doses), care home residence, and recent hospital inpatient stay (5 to 14 days before presentation date) as covariates. Unthresholded *p*-values are reported, allowing comparison of the contribution of each risk factor to prediction of the outcome (the logarithm of the *p*-value scales with the deviance explained). With this incidence density sampling design, the conditional odds ratio is the rate ratio. The reader is cautioned that rate ratios cannot be estimated from unconditional odds ratios because of the matched design [8,9]. For any variable with two or more levels, rate ratios are estimated with respect to a reference category for which the rate ratio is 1. For each drug, the reference category is all those not on the drug. Although individuals prescribed biologics in other centres will be misclassified as not on biologics, this does not seriously affect the estimate of the estimate of the rate ratio because matching on general practice ensures that patients on the biologics list are compared with other individuals from the catchment population of the clinics that provided the biologics list. We have reported both univariate and multivariate rate ratios: for identifying those at high risk, the univariate rate ratios are most relevant, but for inference about possible drug effects the multivariate rate ratios are most relevant.

## Results

The biologics list comprised 4633 individuals: the diagnostic category was rheumatoid arthritis in 2702, psoriatic arthritis or other seronegative arthropathy in 1765, connective tissue disorder in 141, and other conditions in 25. Of the 4633 individuals on the list, 433 had been diagnosed with COVID-19 by 22 November 2021. Of these 433 cases, 58 were hospitalised within 14 days, 7 entered critical care within 21 days and 14 were fatal within 28 days. Of the 4633, 2527 (55%) had been added by PHS to the shielding list based on criteria suggested by the British Society of Rheumatology [10]. Of those added to the shielding list 43 (1.7%) were hospitalised with COVID-19, compared with 15 (0.7%) of those not added. The algorithm used by PHS to identify those eligible for shielding thus discriminated between low-risk and high-risk patients.

Of the 4633 on the biologics list, 2586 were sampled in the REACT-SCOT case-control study. Of these 2586, 2583 had a rheumatology outpatient consultation within the last five years but only 1229 had a dispensed prescription for a conventional synthetic DMARD within the last 240 days. Of the 2586 who were sampled in the case-control study, 1581 had a hospital discharge diagnosis with a rheumatology code.

In comparison with those with no rheumatologic diagnosis, the rate ratio for hospitalized or fatal COVID-19 in those with a hospital discharge diagnosis with a rheumatology code was 2.68 (95% CI 2.47 to 2.91) in those with rheumatoid arthritis, 3.27 (95% CI 2.77 to 3.86) in those with psoriatic or other seronegative arthritis, and 2.28 (95% CI 2.03 to 2.57) in those with connective tissue disorders.

Table 1 shows that the rate ratio for hospitalised COVID-19 associated with DMARD treatment (with those not on DMARDs as reference category) was 2.14 (95% CI 2.02 to 2.26) in those on csDMARDs, 2.01 (95% CI 1.38 to 2.91) in those on TNF inhibitors as the only biologic agent, and 3.83 (95% CI 2.65 to 5.56) in those on other biologic agents.

**Table 1.** Rate ratios for hospitalized or fatal COVID-19 associated with disease-modifying antirheumatic drug treatment

|  | Controls<br>(405597) | Cases (44020) | Univariate |  | Multivariable |  |  |
| --- | --- | --- | --- | --- | --- | --- | --- |
|  |  |  | Rate ratio (95%<br>CI) | *p*-value | Rate ratio (95%<br>CI) | *p*-value |  |
| **Vaccine doses** |  |  |  |  |  |  |  |
| Unvaccinated | 278309 (69%) | 32740 (74%) |  | . |  | . |  |
| 1 dose | 26631 (7%) | 2528 (6%) | 0.51 (0.48, 0.54) | $1 \times 10^{-116}$ | 0.46 (0.43, 0.48) | $3 \times 10^{-142}$ | |
| 2 doses | 96304 (24%) | 8563 (19%) | 0.27 (0.26, 0.28) | $5 \times 10^{-570}$ | 0.25 (0.23, 0.26) | $3 \times 10^{-583}$ | |
| 3 doses | 4353 (1%) | 189 (0%) | 0.07 (0.06, 0.09) | $2 \times 10^{-173}$ | 0.08 (0.06, 0.09) | $6 \times 10^{-154}$ | |
| Care home | 16985 (4%) | 6244 (14%) | 5.3 (5.0, 5.5) | $9 \times 10^{-1383}$ | 5.0 (4.8, 5.3) | $2 \times 10^{-1006}$ | |
| Recent hospital stay | 6602 (2%) | 10131 (23%) | 19.3 (18.6, 20.0) | $3 \times 10^{-5429}$ | 18.5 (17.8, 19.2) | $1 \times 10^{-4951}$ | |
| Any csDMARD | 7017 (2%) | 1565 (4%) | 2.14 (2.02, 2.26) | $4 \times 10^{-153}$ | 2.26 (2.13, 2.41) | $1 \times 10^{-140}$ | |
| **Biologics category** |  |  |  |  |  |  |  |
| No biologic | 405333 (100%) | 43946 (100%) |  | . |  | . |  |
| TNFi only | 164 (0%) | 34 (0%) | 2.01 (1.38, 2.91) | $2 \times 10^{-4}$ | 1.34 (0.88, 2.04) | 0.2 | |
| Other biologic or JAKi | 100 (0%) | 40 (0%) | 3.83 (2.65, 5.56) | $1 \times 10^{-12}$ | 2.87 (1.89, 4.35) | $7 \times 10^{-7}$ | |

Presentation dates up to 22 November 2021
csDMARD exposure is defined as:
(i) any prescription for a conventional synthetic disease-modifying antirheumatic drug in the last 240 days and
(ii) a rheumatology outpatient consultation in the last five years.
Rate ratios are estimated by conditional logistic regression
Multivariable model includes all covariates shown in the table

Table 2 shows the rate ratios for hospitalized or fatal COVID-19 associated with each specific csDMARD or biologic class. The univariate rate ratios associated with conventional synthetic DMARDS ranged from 1.66 (95% CI 1.51 to 1.82) for methotrexate to 5.4 (95% CI 4.4 to 6.7) for prednisolone dose equivalent to more than 10 mg /day. The rate ratio associated with biologics was highest in those on B cell depletion [5.9 (95% CI 3.1 to 11.4) but the confidence intervals were wide and there was no clear evidence that risk varied between different classes of biologic or tsDMARD agent. In a multivariable analysis the effect sizes associated with most drug classes were reduced but the association with glucocorticoids as prednisolone dose equivalent remained strong: rate ratio 5.2 (95% CI 4.1 to 6.6).

**Table 2.** Rate ratios for hospitalized or fatal COVID-19 associated with treatment with specific disease-modifying anti-rheumatic drugs

|  | Controls<br>(405597) | Cases (44020) | Univariate |  | Multivariable |  |
| --- | --- | --- | --- | --- | --- | --- |
|  |  |  | Rate ratio (95%<br>CI) | *p*-value | Rate ratio (95%<br>CI) | *p*-value |
| **Vaccine doses** |  |  |  |  |  |  |
| Unvaccinated | 278309 (69%) | 32740 (74%) |  |  |  |  |
| 1 dose | 26631 (7%) | 2528 (6%) | 0.51 (0.48, 0.54) | $1 \times 10^{-116}$ | 0.46 (0.43, 0.48) | $2 \times 10^{-142}$ |
| 2 doses | 96304 (24%) | 8563 (19%) | 0.27 (0.26, 0.28) | $5 \times 10^{-570}$ | 0.25 (0.24, 0.26) | $2 \times 10^{-581}$ |
| 3 doses | 4353 (1%) | 189 (0%) | 0.07 (0.06, 0.09) | $2 \times 10^{-173}$ | 0.08 (0.06, 0.09) | $9 \times 10^{-153}$ |
| Care home | 16985 (4%) | 6244 (14%) | 5.3 (5.0, 5.5) | $9 \times 10^{-1383}$ | 5.0 (4.8, 5.3) | $1 \times 10^{-1006}$ |
| Recent hospital stay | 6602 (2%) | 10131 (23%) | 19.3 (18.6, 20.0) | $3 \times 10^{-5429}$ | 18.5 (17.8, 19.2) | $5 \times 10^{-4944}$ |
| **Conventional synthetic DMARDs** |  |  |  |  |  |  |
| Methotrexate | 3010 (1%) | 531 (1%) | 1.66 (1.51, 1.82) | $2 \times 10^{-26}$ | 1.29 (1.15, 1.45) | $1 \times 10^{-5}$ |
| Hydroxychloroquine | 1898 (0%) | 361 (1%) | 1.80 (1.61, 2.02) | $4 \times 10^{-24}$ | 1.17 (1.02, 1.35) | 0.02 |
| Sulfasalazine | 1661 (0%) | 420 (1%) | 2.41 (2.16, 2.68) | $5 \times 10^{-57}$ | 2.09 (1.83, 2.38) | $1 \times 10^{-27}$ |
| Leflunomide | 265 (0%) | 52 (0%) | 1.86 (1.38, 2.50) | $5 \times 10^{-5}$ | 1.21 (0.86, 1.71) | 0.3 |
| **Glucocorticoids (prednisolone equivalent mg daily)** |  |  |  |  |  |  |
| 0 | 403211 (99%) | 43214 (98%) |  |  |  |  |
| >0 to 5 | 1574 (0%) | 467 (1%) | 2.78 (2.50, 3.08) | $2 \times 10^{-81}$ | 2.53 (2.24, 2.86) | $3 \times 10^{-51}$ |
| >5 to 10 | 566 (0%) | 201 (0%) | 3.33 (2.83, 3.92) | $7 \times 10^{-48}$ | 2.73 (2.26, 3.30) | $3 \times 10^{-25}$ |
| >10 | 246 (0%) | 138 (0%) | 5.4 (4.4, 6.7) | $9 \times 10^{-56}$ | 5.2 (4.1, 6.6) | $1 \times 10^{-40}$ |
| **Biologics and targeted synthetic DMARDs** |  |  |  |  |  |  |
| TNF inhibitors | 164 (0%) | 34 (0%) | 2.00 (1.38, 2.90) | $2 \times 10^{-4}$ | 1.44 (0.94, 2.20) | 0.09 |
| B cell depletion | 25 (0%) | 15 (0%) | 5.9 (3.1, 11.4) | $7 \times 10^{-8}$ | 3.61 (1.68, 7.77) | 0.001 |
| IL6 inhibitors | 21 (0%) | 9 (0%) | 4.07 (1.86, 8.90) | $4 \times 10^{-4}$ | 3.04 (1.23, 7.51) | 0.02 |
| IL17 inhibitors | 24 (0%) | 5 (0%) | 1.79 (0.66, 4.83) | 0.2 | 0.99 (0.30, 3.22) | 1 |
| JAK inhibitors | 27 (0%) | 12 (0%) | 4.37 (2.20, 8.68) | $2 \times 10^{-5}$ | 2.30 (1.04, 5.07) | 0.04 |

Presentation dates up to 22 November 2021.
For each conventional synthetic DMARD, exposure is defined as:
(i) any prescription for that drug in the last 240 days and
(ii) a rheumatology outpatient consultation in the last five years.
Rate ratios are estimated by conditional logistic regression
Multivariable model includes all covariates shown in the table

## Discussion

### Statement of principal findings

We have shown that:

- Individuals treated with csDMARDs for inflammatory rheumatic diseases have more than two-fold increased risk of hospitalised COVID-19 in comparison with the general population. This increased risk may be at least partly attributable to disease rather than to immunosuppressive therapy.
- The rate ratio for hospitalised COVID-19 in those treated with TNF inhibitors was similar to that in those treated with methotrexate, but the rate ratio in those treated with B-cell depleting agents was higher than in those treated with methotrexate.
- Individuals treated with those treated with glucocorticoids at doses equivalent to more than 10 mg/day prednisolone had a markedly increased risk of hospitalisation with COVID-19

### Strengths and limitations

Strengths of this study are the complete ascertainment of cases, the comprehensive linkage to electronic health records, and the incidence density sampling design that controls for calendar time, age, sex and general practice.

A limitation is that the data on prescribin of biologics is only a snapshot of those on biologics in March 2020, and it covers only about one-third of the Scottish population. Hospital outpatient prescribing is not recorded in electronic form within the NHS, and the prescription records held by the medication homecare services companies were not available for this study. Although the clinics that provided ths biologics list are not representative of the Scottish population, the matching of cases and controls on general practice ensures that the controls are drawn from the catchment population of these clinics. However because the coverage of the list is limited, the sample size is not large enough to estimate reliably the effects of specific classes of biologics such as B cell depleters, or the efficacy of vaccines in those on biologics.

### Comparison with other studies

The rate ratio of 2.1 for hospitalised COVID-19 in rheumatology patients treated with csDMARDs is rather higher than the rate ratio of 1.3 for mortality associated with any diagnosis of rheumatoid arthritis, lupus or psoriasis (not restricted to those with arthritis) in the OpenSAFELY study [11]. The rate ratios associated with immunosuppressive therapy in rheumatology patients are modest in comparison with the rate ratio of 13 that we have reported for severe COVID-19 in solid organ transplant recipients [1].

Others have reported that hospitalised COVID-19 is associated with use of glucocorticoids and B cell depleting agents but not with TNF inhibitors. In a registry of 600 rheumatology patients diagnosed with COVID-19, the odds ratio for hospitalisation (with methotrexate use as reference category) was 2.1 for glucocorticoid use equivalent to > 10 mg/day predisone, and 0.4 for TNF inhibitors [12]. In a later analysis based on 2869 patients with rheumatoid arthritis the odds ratios (with TNF inhibitors as reference category) were 4.5 for rituximab and 2.1 for JAK inhibitors [13]. In a French series of 694 patients the odds ratio for fatal disease was 2.8 for any glucocorticoid, 0.2 for TNF inhibitors and 3.1 for rituximab. [14]. In an Israeli cohort of 6112 psoriasis patients the rate ratio for hospitalisation associated with TNF inhibitors, with methotrexate as reference category, was 0.08 [15].

In this study the rate ratio for hospitalised COVID-19 was similar in users of TNF inhibitors to that in methotrexate users, with no support for a protective effect of TNF inhibitors against severe COVID-19. Other biologics including JAK inhibitors and B cell depleting-therapy were associated with higher risk of severe COVID-19, though the numbers are too small for reliable estimates of the risk associated with each drug class. Rituximab therapy is associated with failure to seroconvert in response to COVID-19 vaccines [16] but the extent to which this affects vaccine efficacy against severe disease is not known. BSR guidance in early 2020 had suggested that hydroxychloroquine and sulfasalazine were unlikely to influence susceptibility to COVID-19 [10], yet the rate ratio for hospitalised COVID-19 was at least as high in those treated with hydroxychloroquine and sulfasalazine as in those treated with methotrexate: a possible explanation for this is that the elevated risk associated with csDMARDs is related to the disease rather than to any immunosuppressive effects of these drugs.

### Clinical implications

This study suggests that all patients on DMARDs are at significantly increased risk of hospitalised COVID-19 compared with the general population. Within the group of treatments that are currently available, methotrexate, HCQ and TNF inhibitors appear to be more favourable than JAK inhibitors and B-cell depleting agents. Prednisolone is associated with high risk even at modest doses, and the risk increases steeply with average dose. A larger study is required to estimate the association of severe COVID-19 with other biologics such as IL-6 and IL-17 inhibitors. Our study brings the risk-benefit of all immunosuppressive therapies for inflammatory rheumatic disease into sharp focus. Most rheumatology patients on immunosuppressant therapy have now received third primary doses of COVID-19 vaccines as recommended; for the effectiveness of this intervention to be monitored it will be necessary to capture and link data on prescribing of csDMARD, biologics and tsDMARDS.

## Supporting information

STROBE checklist for case-control studies

## Data Availability

The component datasets used here are available via the Public Benefits and Privacy Panel for Health and Social Care at https://www.informationgovernance.scot.nhs.uk/pbpphsc/ for researchers who meet the criteria for access to confidential data. All source code used for derivation of variables, statistical analysis and generation of this manuscript is available on https://github.com/pmckeigue/covid-scotland_public.

## Declarations

### Public and Patient Involvement statement

This study was conducted under approvals from the Public Benefit and Privacy Panel for Health and Social Care which includes public and patient representatives.

### Ethics approval and information governance

This study was performed within Public Health Scotland as part of its statutory duty to monitor and investigate public health problems. Under the UK Policy Framework for Health and Social Care Research set out by the NHS Health Research Authority, this does not fall within the definition of research and ethical review is not required. Individual consent is not required for Public Health Scotland staff to process personal data to perform specific tasks in the public interest that fall within its statutory role. The statutory basis for this is set out in Public Health Scotland’s privacy notice.

A Data Protection Impact Assessment (DPIA) allows Public Health Scotland staff to link existing datasets.

### Transparency declaration

PM as the manuscript’s guarantor affirms: that the manuscript is an honest, accurate, and transparent account of the study being reported; that no important aspects of the study have been omitted; and that any discrepancies from the study as originally planned and registered have been explained. This manuscript has been generated directly from the source data by a reproducible research pipeline.

### Contributors

All authors provided substantial contributions to the conception of the study and the drafting of the manuscript. PM undertook the statistical analysis. All authors contributed to revising the manuscript critically for important intellectual content and approved the final manuscript.

## Acknowledgements

We thank Jen Bishop, Ciara Gribben and David Caldwell for undertaking the linkage analysis within Public Health Scotland, and staff from the biologics service in the contributing centres for preparing the list of patients receiving these treatments.

